# Has the COVID-19 Pandemic Changed Parental Attitudes and Beliefs Regarding Vaccinating their Children against the Flu?

**DOI:** 10.1101/2023.05.10.23289801

**Authors:** Liora Shmueli

## Abstract

**Introduction:** This study assessed whether the COVID-19 pandemic has altered parents’ attitudes toward vaccinating their children against the flu, and the contributing socio-demographic, health-related, and behavioral factors, as well as barriers to school-based vaccination programs.

**Methods:** We conducted a cross-sectional online survey of parents of children aged 6 months to 11 years in Israel (n=975) between December 21–31, 2022. A multivariate regression was performed to determine predictors of these parents’ willingness to vaccinate their children aged 6 months to 11 years against the flu in the winter of 2023 (December 2022–February 2023).

**Results:** Overall, 45% of parents stated that they did not intend to vaccinate their children against the flu in the winter of 2023, citing fears of side effects and concerns about vaccine effectiveness. Among those who did not trust the Ministry of Health and pharmaceutical companies prior to the pandemic, this trend increased in 78% of them following the COVID-19 events. In contrast, 39% of parents stated that they had already vaccinated their children against the flu, with an additional 16% intending to do so. Forty-one percent reported an increased intention following the pandemic. Only 37% of parents vaccinated their children at school in grades 2–4, mainly due to a preference for HMO clinics and lack of available nurses at school. The Health Belief Model (HBM) variables, namely, perceived susceptibility, severity and benefits, displayed the largest effect sizes.

**Conclusions:** Understanding the impact of the COVID-19 pandemic on parents’ willingness to vaccinate their children against the flu is crucial. Notably, the pandemic has increased vaccine receptivity among some parents. Healthcare providers and public health officials need to address parents’ concerns about the safety and efficacy of the influenza vaccine to improve vaccination rates among children. Implementing school-based vaccination programs is an important strategy for promoting public health, but may be challenging. To increase uptake, nursing staff in student health facilities should be more accessible, and clear explanations about the efficacy of nasal spray vaccinations should be provided.

## 1. Background

Seasonal influenza, a contagious, infectious viral disease that affects the respiratory system, is caused by the influenza virus, and is mainly common in the fall and winter months. The main symptoms are a runny nose, dry cough, sore throat, fever over 39ºC, headache, and muscle pain. The influenza virus may even cause complications such as secondary infection, in particular pneumonia, hospitalization, and even death. Previous studies have estimated that 99% of deaths in children under 5 years of age with lower respiratory tract infections associated with influenza were in developing countries (Nair et al., 2011; WHO, 2023).

The most effective way and primary approach recommended by the World Health Organization (WHO) and the U.S. Centers for Disease Control and Prevention (CDC) to prevent influenza (flu) is that every person over the age of 6 months should get a flu vaccine every year at the beginning of the flu season. Flu shots are especially recommended for children between the ages of 6 months and 5 years, as they are a high-risk group (CDC, 2022; WHO, 2023). Despite this prioritization, studies conducted before the COVID-19 pandemic found a low degree of flu vaccine compliance in many countries across the globe when it came to child vaccinations. Specifically in the U.S., in the years 2010–2017, only 51–59% of this age group got vaccinated (Gates et al., 2022; Santibanez et al., 2020, pp. 2012–2019), a figure far below the Healthy People 2020 goal of 70% for children (Immunization and Infectious Diseases | Healthy People 2020, n.d.).

This trend continued during the COVID-19 pandemic, one of the most challenging public health crises of the last century. According to the CDC, flu vaccination coverage during the 2021–2022 flu season was 58% among children 6 months through 17 years in the U.S. (Fan et al., 2022). Interestingly, several studies, including systematic reviews, demonstrated a mixed impact of the COVID-19 pandemic on parents’ intention to vaccinate their children against influenza during the pandemic. On the one hand, some studies reported that the pandemic had a positive impact, with parents being more likely to vaccinate their children against influenza, a shift attributed to changes in risk perception due to the COVID-19 pandemic and increased awareness of the importance of vaccination (Goldman et al., 2021). Similarly, Kong’s systematic review of the adult population during the COVID-19 pandemic found a heightened willingness to vaccinate against influenza worldwide (Kong et al., 2022, 2022). On the other hand, other studies reported that the pandemic had a negative impact, with parents being less likely to vaccinate their children due to increased vaccine hesitancy during the COVID-19 pandemic (Day et al., 2022; Humble et al., 2023).

Several factors were found to influence parents’ intention to vaccinate their children against influenza during the COVID-19 pandemic. These include parents’ perception of the severity and risk of COVID-19 and influenza, level of trust in healthcare providers and government recommendations, previous experience with influenza vaccination, and socio-demographic characteristics. Older age, black race, and co-pay have all been associated with decreased influenza vaccine administration (Day et al., 2022).

Only a few studies have investigated caregivers’ hesitancy toward influenza vaccination and assessed the associated factors during the COVID-19 pandemic using the Health Belief Model (HBM) e.g. (Lai et al., 2022). The HBM was widely used in the context of vaccination, particularly influenza vaccination, prior to the COVID-19 pandemic, especially among adults (Bish et al., 2011; Kan & Zhang, 2018; Rosenstock, 1988). This model proposes several factors that explain influenza vaccine acceptance: perceived disease severity; perceived susceptibility; perceived benefits; perceived barriers; and cues to action (Shmueli, 2021, 2023). However, to the best of our knowledge, no studies have examined, using the HBM, the willingness of parents to vaccinate their children against influenza after experiencing the COVID-19 pandemic, during which period some children were vaccinated against COVID-19.

One way to increase flu vaccination rates in this target audience is to administer it in schools. In Israel, as of 2017, flu vaccines are administered in elementary schools through the Ministry of Health’s Student Health Services (in addition to at healthcare maintenance organizations, HMOs), similar to the NHS school-aged immunization service in England. In this preventative, government-funded medical service, students in grades 2–4 are administered nasal spray vaccination by nurses during school hours. Yet, even though this service is provide throughout Israel, many children are not getting vaccinated against the virus (Flu Vaccination at Schools - FAQ, n.d.).

This paper addresses two unmet needs. The first is to assess parental attitude changes to flu vaccinations for their children as a consequence of the COVID-19 pandemic. The second is to identify contributing factors to parental decision-making with regard to vaccinating their children against the flu, in general, including socio-demographic, health-related, and behavioral factors based on the HBM, and with regard to administering flu vaccinations in school, in particular.

## 2. Methods

### Study participants and survey design

We conducted a cross-sectional online survey of 975 Israeli parents to children between the ages of 6 months and 11 years during December 21–31, 2022. The survey was distributed by Sarid Research Institute for Research Services via an online panel containing a wide pool of potential interviewees who had consented to participate in surveys from time to time. Eligible participants were required to be (1) 18+ years old and (2) parents of a child aged 6 months to 11 years. To compose a representative sample of the Jewish adult population in Israel, respondents were sampled by layers based on age, gender, geographic area, and level of religiosity. The questionnaire was pre-tested on a relatively small group of 100 respondents. After the reliability of the questionnaire was verified using a Cronbach α internal reliability test (the HBM section of the questionnaire obtained an internal consistency of Cronbach α=0.85), it was distributed to the rest of the respondents.

### Questionnaire

The questionnaire (Supplementary Questionnaire 1) consisted of 34 mandatory questions, divided into the following sections: (1) Socio-demographic predictor variables; (2) Health-related predictor variables; (3) HBM predictor variables; and (4) Issues related to public health policy (e.g., getting vaccinated at school).

### Variables and measurements

The first dependent variable was *parents’ willingness to vaccinate their children* aged 6 months to 11 years against the flu in the winter of 2023 (December 2020– February 2023), measured in 3 categories: (1) Yes, I have already vaccinated my child; (2) I intent to vaccinate my child in the current winter, and (3) No, I have not vaccinated my child and do not intend to.

The independent variables were grouped into three blocks:

1. Socio-demographic predictor variables: (1) age group; (2) gender; (3) level of education; (4) marital status; (5) socio-economic level^1^; (6) periphery level, determined by the residential area^2^; (7) religiosity level (secular, traditional, religious, orthodox); and (8) working as medical staff.
2. Health-related predictor variables: (1) whether the respondent got vaccinated against COVID-19; (2) whether the respondent already got vaccinated against flu this winter (i.e., around December 2022); (3) whether the children of the respondent got vaccinated against COVID-19; (4) whether a family member suffers from a chronic disease (one or more of the following: heart disease, vascular disease and/or stroke, diabetes mellitus, hypertension, chronic lung disease, including asthma or immune suppression); (5) the existence of past episodes of flu in the current winter; (6) the existence of past episodes of hospitalization in the family in the current winter.
3. HBM predictor variables: (1) perceived susceptibility (included two items); (2) perceived severity (included two items); (3) perceived benefits (included three items); (4) perceived barriers (included one item); (5) cues to action (included three items); and (6) attitude (included one item). The HBM items were measured on a 1–6 scale (1 - strongly disagree; 6 - strongly agree). Negative items were reverse scored. Scores for each item were averaged to generate each of the HBM-independent categories. The Cronbach α internal reliability method revealed the internal consistency of the HBM section to be Cronbach α=0.86 (Supplementary Table 1).

In addition, several issues related to public health policy were examined: (1) public trust in pharmaceutical companies and the Ministry of Health following the COVID-19 pandemic; (2) the main reasons for the reluctancy to vaccinate children against the flu in the winter of 2023; and (3) the main reasons for preferring to vaccinate children in grades 2–4 against flu at the HMO clinic instead of at school (both options are free of charge in Israel).

### Statistical analyses

Data from the online questionnaires were analyzed using SPSS 28 software. To test the reliability of the HBM measures, a Cronbach’s α test was used. To describe the characteristics of the study population, the following methods of descriptive statistics were employed: frequencies, percentages, averages, and standard deviations.

Relationships between dependent and independent variables were examined by univariate analysis. To test the relationship between the demographic variables and willingness to receive a flu vaccine, a series of χ^2^ tests were used. To test the mean differences of the HBM variables at different levels of flu vaccine willingness, we used analysis of variance (ANOVA) tests. Eta squared was used to measure the effect size for significant effects.

To test the multivariate effect of all the significant demographic and health-related variables along with the effects of the HBM variables on willingness to get a flu shot, a multinomial logistic regression analysis was conducted. Only the socio-demographic and health-related variables found to be significant (p<.05) in the univariate analysis were inserted into the regression model as predictors. All the HBM variables were entered into the model as predictors.

## 3. Results

### Participant characteristics

Overall, 975 parents of children aged 6 months to 11 years completed the survey; 52% of them were female (n=505). Almost two thirds of the parents (n=617) were 18– 39 years old. The participants were distributed nearly equally among the three socio-economic categories (low, medium, and high), with 16% of respondents (n=156) living in geographically peripheral regions. Forty percent of the respondents (n=417) were secular, 59% (n=578) held an academic degree, most (89%, n=869) were married, and 25% (n=239) stated having a family member with a chronic disease. The descriptive characteristics of the respondents are provided in Table 1.

**Table 1.** Characteristics of respondents by the intention to vaccinate their children against Flu (n=975)

| | All Subjects | | No Vaccination<br>N=435 (45%) | | Vaccination<br>N=378 (39%) | | Intent to vaccinate<br>N=160 (16%) | | $\chi^2$ | p - value |
| --- | --- | --- | --- | --- | --- | --- | --- | --- | --- | --- |
| **Sociodemographic** | N | % | N | % | N | % | N | % |  |  |
| **Age group** |  |  |  |  |  |  |  |  | 12.49 | **0.002** |
| 18-39 | 617 | 63.3% | 290 | 47.1% | 214 | 34.7% | 112 | 18.2% |  |  |
| 40-60 | 358 | 36.7% | 145 | 40.6% | 164 | 45.9% | 48 | 13.4% |  |  |
| **Gender** |  |  |  |  |  |  |  |  | 10.29 | **0.006** |
| Male | 467 | 48% | 186 | 39.8% | 205 | 43.9% | 76 | 16.3% |  |  |
| Female | 505 | 52% | 247 | 49.1% | 173 | 34.4% | 83 | 16.5% |  |  |
| **Education level** |  |  |  |  |  |  |  |  | 15.46 | **0.02** |
| High school or less | 173 | 17.7% | 81 | 47.1% | 61 | 35.5% | 30 | 17.4% |  |  |
| Non-academic | 224 | 23% | 123 | 54.9% | 72 | 32.1% | 29 | 12.9% |  |  |
| BA | 418 | 42.9% | 167 | 40% | 177 | 42.4% | 73 | 17.5% |  |  |
| MA or higher | 160 | 16.4% | 64 | 40% | 68 | 42.5% | 28 | 17.5% |  |  |
| **Marital status** |  |  |  |  |  |  |  |  | 8.94 | 0.18 |
| Single | 54 | 5.5% | 17 | 31.5% | 29 | 53.7% | 8 | 14.8% |  |  |
| Married | 869 | 89.1% | 392 | 45.2% | 327 | 37.7% | 148 | 17.1% |  |  |
| Divorced | 48 | 4.9% | 24 | 50% | 20 | 41.7% | 4 | 8.3% |  |  |
| Widow | 4 | 0.4% | 2 | 50% | 2 | 50% | 0 | 0% |  |  |
| **Socio-economic level** |  |  |  |  |  |  |  |  | 7.37 | 0.12 |
| Low | 367 | 37.6% | 180 | 49.3% | 130 | 35.6% | 55 | 15.1% |  |  |
| Medium | 303 | 31.1% | 131 | 43.2% | 115 | 38% | 57 | 18.8% |  |  |
| High | 305 | 31.3% | 124 | 40.7% | 133 | 43.6% | 48 | 15.7% |  |  |
| **Peripheral level** |  |  |  |  |  |  |  |  | 2.63 | 0.62 |
| Periphery | 156 | 16% | 71 | 45.5% | 65 | 41.7% | 20 | 12.8% |  |  |
| Intermediate | 399 | 40.9% | 183 | 46% | 150 | 37.7% | 65 | 16.3% |  |  |
| Central | 420 | 43.1% | 181 | 43.2% | 163 | 38.9% | 75 | 17.9% |  |  |
| **Medical Staff** |  |  |  |  |  |  |  |  | 6.41 | **0.04** |
| No | 912 | 93.5% | 416 | 45.7% | 345 | 37.9% | 149 | 16.4% |  |  |
| Yes | 63 | 6.5% | 19 | 30.2% | 33 | 52.4% | 11 | 17.5% |  |  |
| **Religiosity** |  |  |  |  |  |  |  |  | 26.31 | < .001 |
| Secular | 417 | 42.8% | 163 | 39.2% | 190 | 45.7% | 63 | 15.1% |  |  |
| Traditional | 247 | 25.3% | 117 | 47.6% | 86 | 35% | 43 | 17.5% |  |  |
| Religious | 128 | 13.1% | 66 | 51.6% | 36 | 28.1% | 26 | 20.3% |  |  |
| Haredi | 101 | 10.4% | 61 | 60.4% | 26 | 25.7% | 14 | 13.9% |  |  |
| **Health related variables** |  |  |  |  |  |  |  |  |  |  |
| **Chronic Disease** |  |  |  |  |  |  |  |  | 3.6 | 0.17 |
| No chronic disease | 736 | 75.5% | 339 | 46.2% | 282 | 38.4% | 113 | 15.4% |  |  |
| Chronic disease | 239 | 24.5% | 96 | 40.2% | 96 | 40.2% | 47 | 19.7% |  |  |
| **Past episodes of flu current winter parent** |  |  |  |  |  |  |  |  | 3.57 | 0.17 |
| No | 803 | 82.4% | 352 | 43.9% | 310 | 38.7% | 140 | 17.5% |  |  |
| Yes | 172 | 17.6% | 83 | 48.5% | 68 | 39.8% | 20 | 11.7% |  |  |
| **COVID-19 vaccine last year - kids** |  |  |  |  |  |  |  |  | 79.3 | < .001 |
| No | 710 | 73.8% | 365 | 51.6% | 217 | 30.6% | 126 | 17.8% |  |  |
| Yes | 252 | 26.2% | 65 | 25.8% | 157 | 62.3% | 30 | 11.9% |  |  |
| **COVID-19 vaccine - parent** |  |  |  |  |  |  |  |  | 58.17 | < .001 |
| No | 390 | 41.2% | 222 | 57.1% | 105 | 27% | 62 | 15.9% |  |  |
| Yes | 497 | 52.5% | 186 | 37.5% | 232 | 46.8% | 78 | 15.7% |  |  |
| Not yet, but intent to | 60 | 6.3% | 11 | 18.3% | 34 | 56.7% | 15 | 25% |  |  |
| **Past episodes of hospitalization** |  |  |  |  |  |  |  |  | 8.31 | 0.22 |
| No | 872 | 90.6% | 399 | 45.9% | 328 | 37.7% | 143 | 16.4% |  |  |
| Yes, due to flu complications | 4 | 0.4% | 0 | 0% | 4 | 100% | 0 | 0% |  |  |
| Yes, due to COVID-19 complications | 16 | 1.7% | 6 | 37.5% | 8 | 50% | 2 | 12.5% |  |  |
| Yes, other reasons | 70 | 7.3% | 28 | 40% | 29 | 41.4% | 13 | 18.6% |  |  |
| **Flu vaccine current winter - parent** |  |  |  |  |  |  |  |  | 539.36 | < .001 |
| No | 558 | 57.4% | 410 | 73.6% | 95 | 17.1% | 52 | 9.3% |  |  |
| Yes | 224 | 23% | 9 | 4% | 189 | 84.8% | 25 | 11.2% |  |  |
| Not yet, but intent to | 190 | 19.5% | 16 | 8.4% | 92 | 48.4% | 82 | 43.2% |  |  |

A comparison of the sample and the entire population in terms of the characteristics used for sampling (i.e., age, gender, level of religiosity, and geographical area) is provided in Supplementary Table 2.

### Rate of intention to vaccinate children

When parents were asked about their willingness to vaccinate their children aged 6 months to 11 years against the flu in the winter of 2023, 45% (n = 435) stated they had not vaccinated and did not intend to vaccinate their children; 39% (n= 378) responded that they had already vaccinated their children; and the remainder, 16% (n=160), responded that they intend to vaccinate their children. Among those whose children had already been vaccinated or intend to vaccinate, 41% stated that their intention increased following the COVID-19 pandemic.

### Univariate analyses: vaccinate children aged 6 months to 11 years against flu

The results of the univariate analyses between the socio-demographic and health-related variables of parents and their willingness to vaccinate their children aged 6 months to 11 years against the flu are reported in Table 1.

The following socio-demographic variables were found to have a statistically significant correlation (p<0.05) with willingness to vaccinate: age, gender, educational level, medical staff, and religiosity level. The health-related variables found to have a statistically significant correlation (p<0.05) with willingness to vaccinate were: children were vaccinated in the previous year against COVID-19, parent was vaccinated against COVID-19 and/or flu in the current winter.

Specifically, the findings show that women were less likely to vaccinate their offspring compared to men (49% vs. 40%, respectively, χ2 (2)=10.29, p=.006), as were parents under the age of 40 compared to older one (47% vs. 41%, respectively, χ2 (2)=12.49, p=.002), non-academics compared to those with an academic degree (55% vs.40% respectively, χ2 (2)=15.46, p=.02), non-medical occupations compared to medical staff (46% vs. 30%, respectively, χ2 (2)=6.41, p=.04), ultra-orthodox Jews (*haredi*) compared to secular Jews (60% vs. 39%, respectively, χ2 (6)=26.31, p<.001).

Interestingly, parents whose children did not receive a COVID-19 vaccine in the year 2022 expressed significantly higher recalcitrance to vaccinate their children against flu in the winter of 2023 than those whose children did receive a COVID-19 vaccine in the year 2022 (52% vs. 26%, respectively, χ2 (2)=79.3, p<.001).

Parents who did not vaccinate against COVID-19 were more recalcitrance to vaccinate their children aged 6 months to 11 years against the flu compared to those who were vaccinated (57% vs. 38%, respectively, χ2 (4) =58.17, p<.001). Lastly, parents who did not opt to get vaccinated against the flu in the current winter conveyed greater recalcitrance to vaccinate their children aged 6 months to 11 years against the flu compared to those who were vaccinated (74% vs. 4%, respectively, χ2 (4) =539.36, p<.001).

No significant difference was found for the other demographic variables assessed, i.e., socio-economic level, marital status, periphery level, a family member with a chronic disease, past parental episodes of flu in the current winter, and past episodes of hospitalization in the family in the previous year.

The results of the analysis of variance (ANOVA) aimed at testing the mean differences between the HBM variables at the different levels of readiness to vaccinate children aged 6 months to 11 years against flu in the winter of 2023 are reported in Table 2. The perceived susceptibility (F(2,970)=361.7, p<.001, η2=.43), perceived severity (F(2,970)=15.47, p<.001, η2=.03) perceived benefits (F(2,970)=393.47, p<.001, η2=.45) perceived barriers (F(2,970)=94.9, p<.001, η2=.16), cues to action (F(2,970)=281.29, p<.001, η2=.37) and attitude (F(2,970)=53.43, p<.001, η2=.10) all had a statistically significant effect on parents’ willingness to vaccinate their offspring.

**Table 2.** Analysis of variance.

| **Variables** | **No Vaccination** |  | **Vaccination** |  | **Intent to vaccinate** |  | **F-test** | **P value**<br>**(Two-tail)** | **Effect size**<br>**($\eta^2$)** |
| --- | --- | --- | --- | --- | --- | --- | --- | --- | --- |
|  | **Mean** | **SD** | **Mean** | **SD** | **Mean** | **SD** |  |  |  |
| Susceptibility | 2.50 | 1.13 | 4.49 | 1.12 | 4.16 | 0.94 | 361.70 | <.001 | 0.43 |
| Severity | 2.42 | 1.12 | 2.77 | 0.95 | 2.83 | 0.92 | 15.47 | <.001 | 0.03 |
| Benefits | 2.57 | 1.03 | 4.52 | 1.03 | 4.15 | 1.00 | 393.47 | <.001 | 0.45 |
| Barriers | 4.58 | 1.33 | 3.31 | 1.35 | 3.68 | 1.32 | 94.9 | <.001 | 0.16 |
| Cues to action | 2.37 | 1.18 | 4.18 | 1.12 | 4.00 | 1.11 | 281.29 | <.001 | 0.37 |
| Attitude | 4.03 | 1.65 | 4.97 | 1.20 | 4.95 | 1.08 | 53.43 | <.001 | 0.10 |

### Multivariate analyses: predictors of willingness to vaccinate children against flu

We performed a multinomial logistic regression analysis of the multivariate effect of all the significant demographic and health-related variables along with those of the HBM variables on the willingness of parents to vaccinate their children aged 6 months to 11 years against the flu. The regression analysis accounted for an estimated 71% of the explained variance in the willingness to vaccinate children aged 6 months to 11 years against the flu in the winter of 2023 (Nagelkerke Pseudo R^2^=.71) (see full regression coefficients in Table 3). While none of the socio-demographic variables were significantly associated with the willingness to vaccinate children aged 6 months to 11 years against the flu, four health-related variables were found to be significant predictors. Specifically, an examination of the regression coefficients indicated that when all other variables are controlled, parents who vaccinated their children against COVID-19 in the previous year were 2.52 times more likely to vaccinate their children against the flu in the winter of 2023 than those who did not vaccinate their children against COVID-19 in the previous year (OR=2.52, 95% CI. 1.28–4.95, p=.01). Yet, parents who themselves got vaccinated against COVID-19 were 0.57 times less likely to vaccinate their children for flu in comparison to parents who did not opt for a COVID-19 vaccine (OR=0.57, 95% CI. 0.35– 0.93, p=.03). In the case of parents who got a flu shot in the winter of 2023, they were 24.37 times more likely to vaccinate their children against the flu and 6.25 times more likely to intend to vaccinate their children against the flu in comparison to those who did not get a flu shot that winter (OR=24.37, 95% CI. 8.28–71.74, p<.001; OR=6.25, 95% CI. 1.92–20.29, p<.001). Parents who intended to get a flu shot were 9.3 more likely to vaccinate their children against the flu and 16.46 times more likely to intend to vaccinate their children against the flu in the same winter in comparison to those who did not get a flu shot (OR=9.3, 95% CI. 4.31–20.07, p<.001, OR=16.46, 95% CI. 7.59–35.69, p<.001).

**Table 3.** Multinomial regression- predictors of intention to vaccinate children aged 0.5-11 years with Flu (N=975).

|  | Vaccination |  |  |  |  |  | Intent to vaccinate |  |  |  |  |  |
| --- | --- | --- | --- | --- | --- | --- | --- | --- | --- | --- | --- | --- |
|  | B | SE | P | OR | LLCI | ULCI | B | SE | P | OR | LLCI | ULCI |
| Intercept | -7.57 | 1.21 | 0.00 |  |  |  | -7.04 | 1.25 | 0.00 |  |  |  |
| Age Group (40+) | 0.23 | 0.30 | 0.45 | 1.25 | 0.70 | 2.26 | 0.05 | 0.32 | 0.86 | 1.06 | 0.57 | 1.96 |
| Sex (Male) | -0.17 | 0.27 | 0.53 | 0.84 | 0.49 | 1.44 | 0.14 | 0.29 | 0.62 | 1.16 | 0.66 | 2.03 |
| Education (non academic) | -0.22 | 0.43 | 0.61 | 0.80 | 0.34 | 1.86 | -0.59 | 0.45 | 0.19 | 0.55 | 0.23 | 1.34 |
| Education (BA) | 0.06 | 0.40 | 0.87 | 1.07 | 0.49 | 2.32 | -0.13 | 0.40 | 0.74 | 0.87 | 0.40 | 1.92 |
| Education (MA or higher) | -0.33 | 0.47 | 0.48 | 0.72 | 0.29 | 1.80 | -0.25 | 0.48 | 0.60 | 0.78 | 0.31 | 1.98 |
| Medical staff | -0.25 | 0.61 | 0.68 | 0.78 | 0.24 | 2.56 | -0.34 | 0.63 | 0.58 | 0.71 | 0.21 | 2.43 |
| Religiosity (Traditional) | -0.02 | 0.32 | 0.94 | 0.98 | 0.52 | 1.85 | -0.06 | 0.35 | 0.86 | 0.94 | 0.48 | 1.85 |
| Religiosity (Religious) | -0.65 | 0.39 | 0.09 | 0.52 | 0.24 | 1.11 | -0.11 | 0.39 | 0.78 | 0.90 | 0.42 | 1.94 |
| Religiosity (Haredi) | -0.18 | 0.46 | 0.70 | 0.84 | 0.34 | 2.07 | 0.02 | 0.49 | 0.96 | 1.02 | 0.40 | 2.65 |
| Covid-19 vaccine last year - kids | 0.92 | 0.34 | **0.01** | 2.52 | 1.28 | 4.95 | 0.16 | 0.38 | 0.67 | 1.17 | 0.56 | 2.46 |
| Covid-19 vaccine last year - parent | -0.46 | 0.24 | **0.05** | 0.63 | 0.40 | 1.00 | -0.55 | 0.25 | **0.03** | 0.57 | 0.35 | 0.93 |
| Current flu vaccine-parent | 3.19 | 0.55 | **<.001** | 24.37 | 8.28 | 71.74 | 1.83 | 0.60 | **<.001** | 6.25 | 1.92 | 20.29 |
| Positive Intention flu vaccine-parent | 2.23 | 0.39 | **<.001** | 9.30 | 4.31 | 20.07 | 2.80 | 0.39 | **<.001** | 16.46 | 7.59 | 35.69 |
| **Health Belief Model** |  |  |  |  |  |  |  |  |  |  |  |  |
| Susceptibility | 0.63 | 0.18 | **<.001** | 1.88 | 1.33 | 2.67 | 0.63 | 0.19 | **<.001** | 1.89 | 1.31 | 2.71 |
| Severity | 0.60 | 0.16 | **<.001** | 1.82 | 1.33 | 2.49 | 0.58 | 0.16 | **<.001** | 1.79 | 1.30 | 2.47 |
| Benefits | 0.98 | 0.19 | **<.001** | 2.65 | 1.82 | 3.87 | 0.62 | 0.20 | **<.001** | 1.86 | 1.27 | 2.73 |
| Barriers | -0.29 | 0.11 | **0.01** | 0.75 | 0.61 | 0.92 | -0.30 | 0.11 | **0.01** | 0.74 | 0.59 | 0.92 |
| Cues to action | 0.37 | 0.14 | **0.01** | 1.45 | 1.10 | 1.91 | 0.32 | 0.15 | **0.03** | 1.37 | 1.03 | 1.83 |
| Attitude | -0.15 | 0.12 | 0.21 | 0.86 | 0.68 | 1.09 | 0.05 | 0.13 | 0.73 | 1.05 | 0.81 | 1.35 |

All the HBM tested variables were found to be significant predictors of the willingness of parents to vaccinate their children aged 6 months to 11 years against the flu. Specifically, for each unit increase in susceptibility, the odds of vaccinating increased by 1.88-fold, similar to the odds of intending to vaccinate by 1.89-fold (p<.001). For each unit increase in severity, the odds of vaccinating increased by 1.82-fold (p<.001). For each unit increase in benefits, the odds of vaccinating increased by 2.65-fold, and the odds of intending to vaccinate by 1.86-fold (p<.001). For each unit increase in barriers, the odds of vaccinating decreased by .75-fold, and the odds of intending to vaccinate by .74-fold (p=.01). For each unit increase in cues to action, the odds of vaccinating increased by 1.45-fold and the odds of intending to vaccinate by 1.37-fold (p=.01). A complete description of the model, goodness of fit indices and regression coefficients are presented in Table 3.

### Main reasons for the reluctancy to vaccinate children against the flu in the winter of 2023

Forty five percent (n=435) of the parents did not vaccinate and did not intend to vaccinate their children against the flu in the winter of 2023. The most common reasons for this decision were fear of side-effects of the vaccine (28%) and concerns about vaccine effectiveness (22%). Other reasons were a lack of trust in the Ministry of Health (13%) or in pharmaceutical companies and vaccines (13%); of these 169 parents, 132 (78%) stated that the lack of trust increased following the COVID-19 events. Another stated reason was the impression that natural vaccination affords greater protection than an injected vaccine (17%) (see Supplementary Table 3 for the complete list of reasons).

### The main reasons for preferring to vaccinate children in grades 2–4 against the flu at a HMO instead of at the school

The main reasons that parents opted to vaccinate their children in grades 2–4 at a HMO clinic rather than through the Student Health Services at the elementary school during school hours with a nasal spray (n=188) were: preference to vaccinate in the presence of the parent (36%), no nurse availability, so there were no vaccination campaign at the school (23%), wanted to vaccinate the children at school, but missed the vaccination day at school (16%), and considering the nasal spray to be not effective, whereas the HMO clinics administered flu shots, perceived by the parent to be more effective (12%).

## 4. Discussion and Conclusion

### Discussion

In this study, we examined parents’ willingness to vaccinate their children aged 6 months to 11 years against the flu following the COVID-19 pandemic, in the winter of 2023, and to identify contributing factors to parental decision-making in this regard, including socio-demographic, health-related, and behavioral factors. In addition, we explored parents’ attitudes toward vaccinating their children in school settings as part of Student Health Services.

Previous research has examined parents’ attitudes towards vaccinating their children against the flu, although only a few studies have explored this in the context of the COVID-19 pandemic. The present study found that almost half of the parents who participated in the surveyed did not intend to vaccinate their children against the flu in the winter of 2023. Of the remaining 55%, 39% had already vaccinated their kids, and 16% intended to do so. These findings are consistent with a recent study conducted at the end of 2022 in 14 Eastern Mediterranean region (EMR) countries, which demonstrated that half of parents in these countries were hesitant about vaccinating their children against influenza (Fadl et al., 2023). It is also in line with a study conducted among Chinese middle-school students, which found an overall vaccination rate of 38.2% in this group (Asihaer et al., 2022).

The socio-demographic characteristics we found to be associated with parents’ willingness to vaccinate their children against the flu were, too, in line with previous studies. These factors include male gender (Kini et al., 2022), older parents (Li et al., 2022), higher levels of education (Li et al., 2022), and a medical profession (Sallam et al., 2022). These findings underscore the need to consider socio-demographic factors when designing interventions and campaigns to increase vaccine uptake.

In the hierarchical regression analysis, the HBM variables perceived susceptibility, perceived severity, and perceived benefits displayed the largest effect sizes. This finding highlights these constructs’ importance as predictors of vaccine acceptance and their utility as targets for interventions aimed at promoting health behaviors.

Understanding the impact of the COVID-19 pandemic on parents’ willingness to vaccinate their children against the flu post-pandemic is crucial. As our findings indicate, the pandemic has led to greater vaccine receptivity among some parents. Notably, approximately 41% of parents whose children have already been vaccinated or are planning to get vaccinated reported an increase in their intention to vaccinate against the flu following the pandemic. These findings are consistent with a systematic review that highlighted a rise in the intention to vaccinate against the flu during the pandemic (Kong et al., 2022). Thus, the pandemic has provided a window of opportunity to promote influenza vaccination and decrease vaccine hesitancy among parents. Encouragingly, the increased awareness as to the importance of vaccination due to the COVID-19 pandemic could lead to improved vaccination rates and better protection against both influenza and COVID-19.

While the pandemic has increased awareness to the importance of vaccination, it has also heightened vaccine hesitancy among some parents, particularly those skeptical about vaccines in general. Most of the parental concerns raised in this study center around vaccine safety and efficacy issues. As has been previously reported, vaccine safety and potential side-effects are the main barriers to vaccination (Asihaer et al., 2022; Price et al., 2022). Interestingly, lack of trust in the Ministry of Health and in the pharmaceutical companies increased among 78% of the vaccine skeptics in our study following the COVID-19 events. This lack of trust can arise from a range of factors, such as misinformation, lack of transparency and conspiracy theories about vaccines, all of which contribute to confusion and mistrust among some parents. It is, therefore, important that healthcare providers and public health officials address these concerns and provide accurate information about the safety and efficacy of the influenza vaccine and work towards rebuilding trust in the Ministry of Health to help increase the vaccination rate among parents and children.

Our results further indicate that only 37% of parents to kids in grades 2–4 chose to vaccinate their children at school, mainly because they preferred doing so at the HMO clinic in their presence or due to the unavailability of a nurse to administer the vaccine at the school. The lack of personnel at the school may lead parents to opt for alternative vaccination options, such as visiting the HMO clinic. Overall, while implementing school-based vaccination programs may be challenging, it is an important strategy for improving vaccination rates and promoting public health. Indeed, a recent study found that sending a behaviorally informed invitation letter and reminders can increase childhood influenza vaccinations through schools and general practitioner (GP) practices (Howell-Jones et al., 2023).

Nevertheless, it is important to recognize this study’s limitations when interpreting the reported results. First, it is important to be aware of the predictive limitation of a cross-sectional study. Namely, since the exposure and outcome are simultaneously assessed, it is not possible to determine a temporal or causal relationship between them, Therefore, it cannot be ruled out that other variables created pseudo-correlations. Moreover, the study used self-report of influenza vaccine acceptance and intention to influenza vaccine in the coming winter, but the self-report of actual behavior may be biased, unlike monitoring actual vaccination. Finally, the study used a cross-sectional observational design that does not allow one to derive any causal conclusions.

### Conclusions

Understanding the impact of the COVID-19 pandemic on parents’ willingness to vaccinate their children against the flu is crucial. Notably, the pandemic has led to enhanced vaccine receptivity among some parents. Healthcare providers and public health officials should address parents’ concerns about the safety and efficacy of the influenza vaccine as a means to improve vaccination rates among children. Implementing school-based vaccination programs is an important strategy for promoting public health, but may be challenging. To increase uptake, nursing staff in student health facilities should be more accessible, and clear explanations about the efficacy of nasal spray vaccinations should be provided.

## Data Availability

All data produced in the present study are available upon reasonable request to the authors

## List of abbreviations

HBM: Health Belief Model
WHO: World Health Organization
CDC: The U.S. Centers for Disease Control and Prevention

## Declarations

### Ethics approval and consent to participate

The study was approved by the Ethics Committee for Non-clinical Studies at Bar-Ilan University, Israel. The ethics form was signed by the committee head on December 8, 2022. Informed consent for participating in the study was obtained digitally from all subjects through the online questionnaire distributed by Sarid Research Institute. All methods were carried out in accordance with the relevant guidelines and regulations of the Ethics Committee for Non-clinical Studies at Bar Ilan-University, Israel.

Specifically, at the beginning of the questionnaire, participants were asked whether they agree to participate in the research and whether they are 18 years old or older. Only participants who answered these two questions positively were allowed to continue with the questionnaire. Participants were also informed that their participation was voluntary and that they had the right to leave at any time without providing any explanation. Participants in the panel are rewarded (by Sarid Institute) for participating in surveys from time to time, according to the estimated time it takes them to complete a survey. In our case, each participant who completed the questionnaire received 1.8 NIS.

### Consent for publication

Not applicable

### Availability of data and material

All data generated or analyzed during this study are included in this published article [and its supplementary information files].

### Authors’ contributions

LS was responsible for the study conception and design, data collection and analysis, and writing the manuscript.

## Acknowledgements

Not applicable

## Author information

Liora Shmueli is a faculty member at the Department of Management, Bar-Ilan University.

## Supplementary Tables

**Supplementary Table 1.** Items and internal consistency for assessing measures of the HBM.

| Measures | Items | $\alpha$ |
| --- | --- | --- |
| Perceived susceptibility | I believe that if I my children will get vaccinated, the likelihood of them getting infected with Flu will decrease | 0.88 |
|  | I believe that if my children will not get vaccinated, the likelihood of our family and relatives getting infected with Flu will increase |  |
| *Perceived severity | Even if my children will get infected with Flu I do not think it will cause them significant suffering or complications | 0.64 |
|  | Even if my children will get infected with Flu, the likelihood of them recovering from the disease is very high |  |
| Perceived Benefits | I believe that the Flu vaccine will be highly effective in preventing my children from experiencing significant complications of the disease | 0.92 |
|  | I believe that if my children will get vaccinated against Flu, the likelihood that they will miss school days (school / kindergarten) will decrease |  |
|  | I believe that if my children will get vaccinated against Flu, the likelihood of us (the parents) losing work days will decrease |  |
| Perceived barriers | I am afraid that the Flu vaccine has side effects | - |
| Cues to action | The likelihood of vaccinating my children against Flu will increase if the health minister will express their support for the benefits of vaccinating children | 0.88 |
|  | The likelihood of vaccinating my children against Flu will increase if my pediatrician will recommend that they be vaccinated |  |
|  | The likelihood of vaccinating my children against Flu will increase if the vaccine will be administered by the educational system |  |
| Attitude | I support vaccine and My children are vaccinated according to the standard immunization schedule | - |

$\alpha$ Cronbach indicates the internal consistency: **HBM** $\alpha = 0.86$
Items response scale: 1-6 agreement
\* Negative items were reverse scored

**Supplementary Table 2:** Comparing the sample and the entire population in terms of the characteristics used for sampling: age, gender, level of religiosity, and geographical area.

| **Sociodemographic** | **Population** | **sample**<br>**(n=1012)** |  |
| --- | --- | --- | --- |
|  | % | N | % |
| **Age group \*** |  |  |  |
| 18-39 | 58% | 617 | 63% |
| 40-60 | 42% | 358 | 37% |
| **Gender \*** |  |  |  |
| Male | 50% | 467 | 48% |
| Female | 50% | 505 | 52% |
| **Level of religiosity \*\*** |  |  |  |
| Secular | 45% | 417 | 43% |
| Traditional | 25% | 247 | 25% |
| Religious | 16% | 128 | 13% |
| Haredi | 14% | 101 | 10% |
| **Geographical area \*\*** |  |  |  |
| *Jerusalem District* | 11% | 93 | 9% |
| *Northern District* | 9% | 124 | 12% |
| *Haifa District* | 11% | 159 | 16% |
| *Central District* | 29% | 271 | 27% |
| *Tel Aviv District* | 20% | 168 | 17% |
| *Southern District* | 14% | 146 | 14% |
| *Judea and Samaria Area* | 6% | 48 | 5% |

\* Jewish population, ages 18-60 (n=3.4 million)
\*\* Jewish population, all ages (n=6.4 million)

**Supplementary Table 3:** Reasons for the reluctancy to vaccinate children against the flu (n = 967 reasons provided by the 435 parents didn’t vaccinate and don’t intend to vaccinate their children against the flu in the winter of 2023

| **Reasons for reluctance to vaccinate children against the flu** | **n** | **%** |
| --- | --- | --- |
| **I am afraid of the side effects of the vaccine** | 272 | 28% |
| I believe that the vaccine is not effective | 212 | 22% |
| The natural vaccine protects more than an injected vaccine | 164 | 17% |
| Lack of trust in the Ministry of Health | 128 | 13% |
| Lack of trust in pharmaceutical companies and vaccines | 128 | 13% |
| I am against all types of vaccines in general | 29 | 3% |
| Inaccessibility | 21 | 2% |
| Medical contraindication (sensitivity to the vaccine) | 13 | 1% |

Respondents could provide more than one reason for the reluctance to vaccinate children against the flu in the winter of 2023. Hence, the number of reasons is larger than the number of respondents

## Footnotes

1 The socioeconomic level according to the Central Bureau of Statistics. It is based on the average gross income for a family in Israel, which is about 19,300 NIS per month when both spouses are employed and about 9,000 NIS per month for a single person, grouped into three levels: low, medium and high.

2 The ‘periphery’ level according to the Israeli Central Bureau of Statistics. It is based on a peripheral index that combines two components: potential accessibility index of the local authority and proximity of the local authority to the boundary of the Tel Aviv district (www.cbs.gov.il). The peripheral index includes local authorities that were classified into 10 clusters. In this study, these clusters were grouped based on their periphery distribution scale into three groups: peripheral, intermediate, and central.

